# COVID-19 pandemic: Power law spread and flattening of the curve

**DOI:** 10.1101/2020.04.02.20051680

**Authors:** Mahendra K. Verma, Ali Asad, Soumyadeep Chatterjee

## Abstract

In this paper, we analyze the real-time infection data of COVID-19 epidemic for nine nations. Our analysis is up to 7 April 2020. For China and South Korea, who have already flattened their infection curves, the number of infected individuals (*I*(*t*)) exhibits power-law behavior before flattening of the curve. Italy has transitioned to the power-law regime for some time. For the other six nations—USA, Spain, Germany, France, Japan, and India—a power-law regime is beginning to appear after exponential growth. We argue that the transition from an exponential regime to a power-law regime may act as an indicator for flattening of the epidemic curve. We also argue that long-term community transmission and/or the transmission by asymptomatic carriers traveling long distances may be inducing the power-law growth of the epidemic.

## 1 Introduction

COVID-19 pandemic has caused major disruptions in the world. As of 1 April 2020, it has infected approximately 1.7 million humans, killed more than 0.1 million individuals, and has brought most of the world to a standstill in lockdowns [1, 2]. Hence, modeling and forecast of this epidemic is of critical importance. In this paper we analyze the publicly available data set given in the *worldOmeter* [1]. The data show that countries that have achieved flattening of the epidemic curve exhibit power law growth before saturation. This feature can be used as an important diagnostic for flattening of the epidemic curve.

Epidemiologists have made various models for understanding and forecasting epidemics. Kermack and McKendrick [3] constructed one of the first models, called SIR model, for epidemic evolution. Here, the variables *S* and *I* describe respectively the numbers of susceptible and infected individuals. The variable *R* represents the removed individuals who have either recovered or died. The SIR model has been generalized to SEIR model that includes *exposed* individuals, *E*, who are infected but not yet infectious [4, 5].

SARS-CoV-2 is an extremely contagious virus. In addition, many infected individuals, called *asymptomatic carriers*, who show mild or no symptoms of infection have contributed significantly to the spread of the epidemic unwittingly [6]. Hence, modelling COVD-19 requires more complex models of epidemiology, including features of quarantine, lockdowns, stochasticity, interactions among population pockets, etc. Note that quarantines and lockdowns help in suppressing the maximum number of infected individuals; such steps are critical for the epidemic management with limited public health resources. The saturation or flattening of the curve in China is attributed to strong lockdowns.

For COVID-19 epidemic, some of the new models have managed to provide good forecasts that appears to match with the data. Peng et al. [7] constructed a seven-variable model (including quarantined and death variables) for epidemic spread in China and predicted that the daily count of exposed and infectious individuals will be negligible by 30 March 2020. Their predictions are in good agreement with the present data. Chinazzi et al. [8] studied the effects of travel restrictions on the spread of COVID-19 in China and in the world, and Hollewell et al. [9] performed feasibility studies of controlling COVID-19 epidemic by isolation. Mandal et al. [10] constructed a India-specific model for devising intervention strategies; they focussed on four metros— Delhi, Mumbai, Kolkata, and Bengaluru—along with intercity connectivity. To account for spatio-temporal behavour, Min et al. [11] simulated how a disease could spread within a network with different mixing styles, and showed that the average epidemic size and speed depend critically on network parameters. In addition, there are many epidemic models that are inspired by population growth models [5, 12]. There are several other models designed to understand the spread of COVID-19 [13–15].

## 2 Data Analysis and Results

In this paper we report our results based on a comprehensive data analysis of nine major countries–China, USA, Italy, France, Spain, Germany, South Korea, Japan and India. We chose the above nations because of the large numbers of positive cases here. For our analysis, we employed the real-time data available at *worldOmeter* [1].

We digitized the data up to 7 April 2020 and studied the temporal evolution of the cumulative count of infected individuals, which is denoted by *I*(*t*), where *t* is time in days. In Fig. 1 we plot the time series of *I*(*t*) and its derivative *İ*(*t*) in semi-logy format using red and blue curves respectively. The derivatives *İ*(*t*) have been computed using Python’s *gradient* function. In these plots we represent time using dates with month/date format; the starting date for each plot is chosen as *t* = 0.

**Fig. 1.**
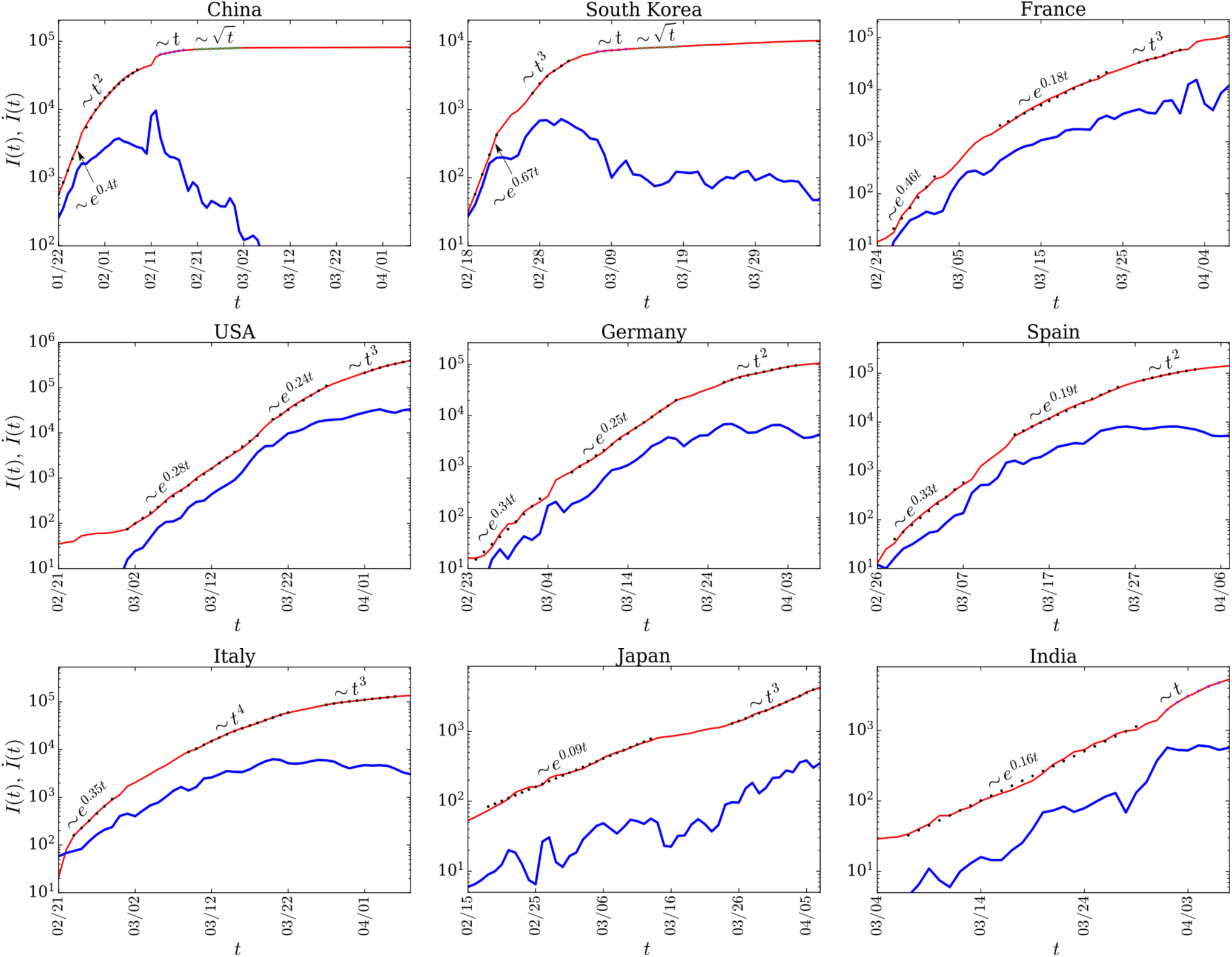
(color online) For the COVID-19 epidemic, the *semi-logy* plots of total infected individuals (*I*(*t*)) vs. time (*t*) (red thin curves) for the nine nations. We also plot *İ*(*t*) vs. *t* (blue thick curves). The black dotted curves represent the best fit curves. For the best fit functions, refer to Table 1.

No single function appears to fit with *I*(*t*), hence we employ different functions to fit at different time intervals. As is well-known, in the early phase, the growth is exponential, but it transitions to power laws subsequently. Hence, we employ exponential function and polynomials for constructing best fit curves to different parts of *I*(*t*). The best functions are listed in Table 1, as well as exhibited in all the plots. We have computed the best fit curves using Python’s *polyfit* function, and the errors as the relative errors between the original data and the fitted data.

**Table 1.**
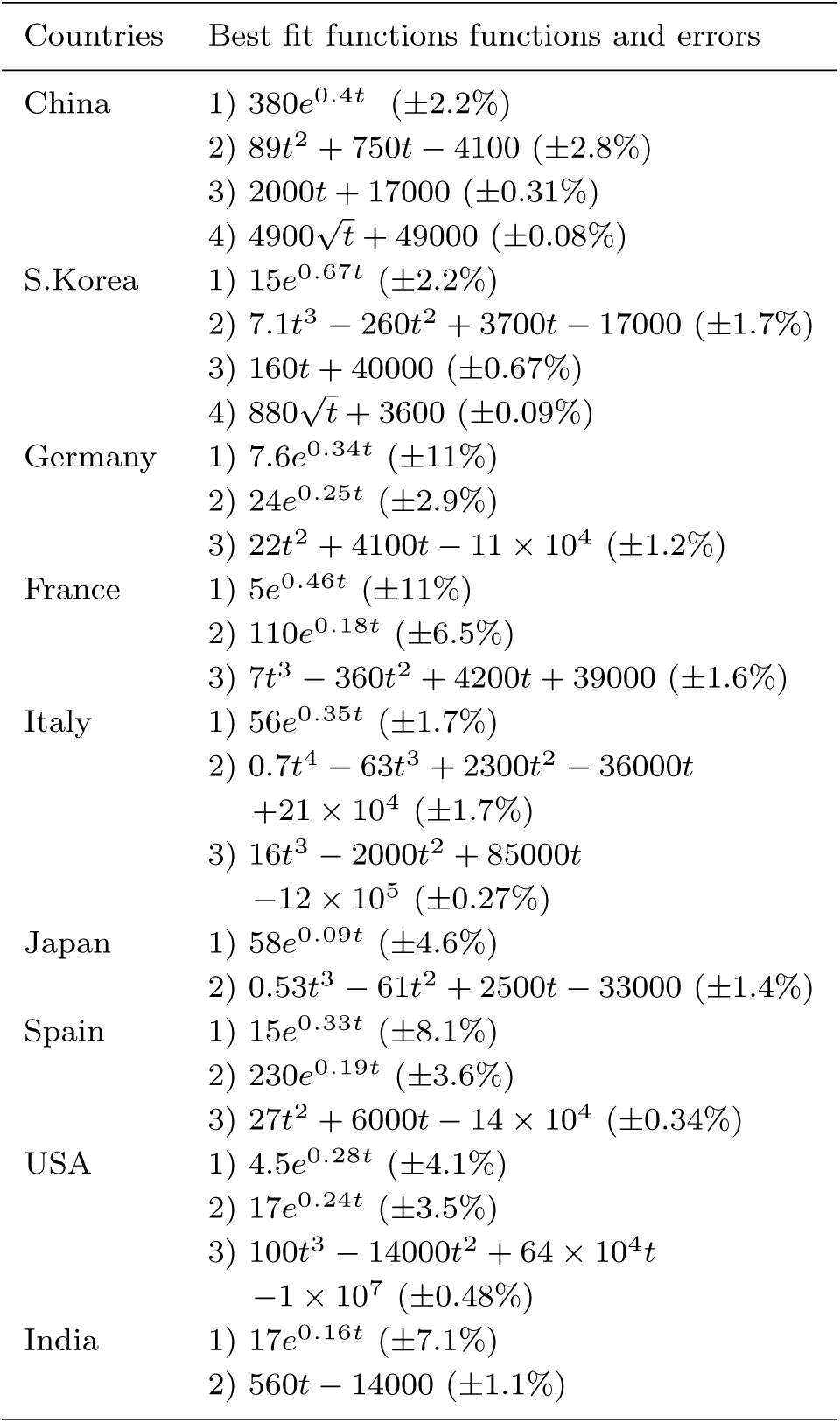
For the COVID-19 data for various countries, the best fit functions and the respective relative errors for various stages of evolution shown in Fig. 1. The figure also exhibits the respective best fit curves.

In the early phase, all the curves exhibit exponential growth as *I*(*t*) = *A* exp(*βt*), where *β* is the growth rate. Interestingly, the *I*(*t*) plots for USA, Spain, Germany, and France require two exponential functions for the fits. For example, Germany’s data requires two functions, exp(0.34*t*) and exp(0.25*t*). Note that the growth rate *β* varies for different countries, which is because *β* depends on various factors such as immunity level of the population, climate, local policy decisions (lockdown, social distancing), etc.

Larger the *β*, larger the growth rate for the infection. Also, the inverse of the constant *β* yields the growth time scale. In fact, in the exponential phase, the number of cases double in time *T* = (log 2)*/β*. For South Korea, *β* = 0.67, hence, *T ≈* 1; that is, *I*(*t*) for South Korea doubled every day in the early phase (18 February to 23 February). The doubling time for India in the exponential phase was log(2)*/*0.16 *≈* 4.3 days. Note that for the exponential regime, *İ ≈ βI*.

After the exponential phase, the curves transition to power laws, polynomials to be more precise. The curve for China exhibits three power laws: *t*^2^, *t*, and then 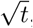, after which *I*(*t*) saturates. For South Korea, we observe *t*^3^ growth rather than *t*^2^ growth. Among the nine nations, only China and South Korea have flattened their epidemic curves. We make a remark that the best fit curves are polynomials (see Table 1); the power laws mentioned above and those indicated in the plots are the leading-order terms of the respective polynomials.

The other seven nations exhibit transition to their power law regimes after the initial exponential growth, with the power law exponents ranging from 2 to 4. Note however that the power law regimes are quite small, except for Italy. We believe that the curves will flatten further down from here, that is, they will transition to subsequent power laws with lower exponents. However, we need more data of subsequent days for a definitive conclusion.

Figure 1 also contains plots for *İ*, derivatives of *I*(*t*), that represents daily new count of infections. Similar to *I*(*t*), *İ* increases exponentially in the beginning. After this, we observe a transition to power law regimes. For the power law *I*(*t*) *∼ Bt*^*n*^, we derive that *İ ∼ I*^1*−*1*/n*^. Clearly, this slope is suppressed compared to the exponential regime by a factor of *I*^*−*1*/n*^. From time *t*_0_, *I*(*t*) doubles at *t* = 2^1*/n*^*t*_0_. For South Korea, *n* = 3, hence for *t*_0_ = 10, the count doubles at *t* = 10 *×* 2^1*/*3^ *≈* 12.6 day, or in the interval of 2.6 days. This is a slower doubling rate than that in the exponential phase, which was one day. Note however that the epidemic growth in the power law regime is still very significant because *I*(*t*) is large. For large *n* (e.g., 4 or 5), *İ ∝ I*, which is same as the formula for the exponential growth (refer to the *t*_4_ regime of Italy and Japan). Also note that in the linear regime, *İ* is constant, implying a constant number of new cases every day.

In Fig. 1, in the exponential regimes, *I*(*t*) and *İ*(*t*) curves run almost parallel to each other because *İ βI*. In contrast, in the power law regimes, *İ* (*t*) deviates from being parallel to *I*(*t*), consistent with the suppression in *İ* (*t*) mentioned in the previous paragraph. For Japan, *İ* exhibits a marginal deviation from the form *İ ≈ βI*(*t*); this is due to the fact that n = 4, which is large.

We can combine the above ingredients into a comprehensive picture for the epidemic forecast, specially for flattening or saturating the *I*(*t*) curve that is prime objective for most affected nations. As illustrated in the schematic diagram of Fig. 2, the *I*(*t*) curve follows four stages: exp(*βt*), *t*^*n*^, *t*, and constant, which are represented by S1, S2, S3, and S4 respectively. It is an elementary observation that the *I*(*t*) curve transitions from a convex form (S1) to a concave form (S2, S3, S4). Such a simple observation of the data reveals insights into the temporal evolution of the epidemic. For example, before flattening of *I*(*t*), we look for flattening of the growth rate *İ*(*t*), which is the third stage in Fig. 2.

**Fig. 2.**
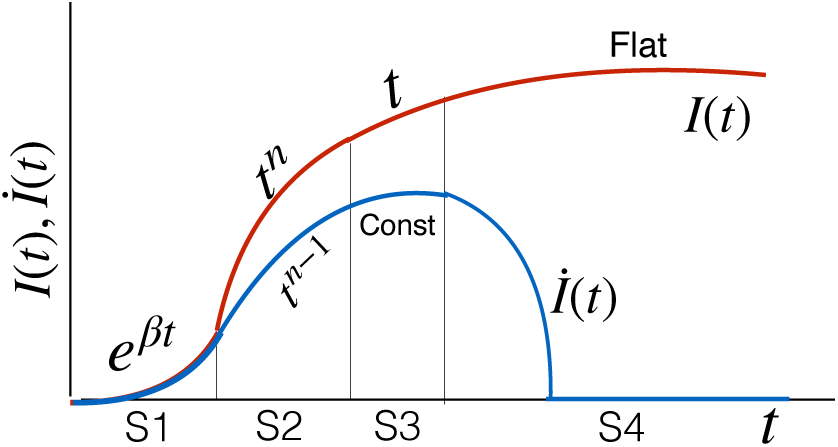
(color online) For COVID-19 epidemic: Schematic plots for *I*(*t*) and its derivative *İ* (*t*) vs. *t*. S1, S2, S3, S4 represent the four stages of the epidemic: exponential growth in count (exp(*βt*)), power law growth (*t*^*n*^), linear growth (*t*), and flat.

Note that the above features of Fig. 2 appear in almost all epidemic models, however with shorter or negligible power law regime. For example, Wu et al. [12] considered a model with *İ* = *rI*^*p*^(1 *−* (*I/κ*)^*α*^), where *r, p, κ, α* are free parameters, to provide a fit to the epidemic curve for China. In our paper, the focus is on the data itself, rather than models that may involve many parameters.

## 3. Discussions and Conclusions

COVID-19 appears to be a unique epidemic to exhibit a power law regime, which was not exhibited by earlier epidemics, such as SARS and EBOLA [16, 17]. We believe this feature to be related to the super spreading of COVID-19 by asymptomatic carriers. As is evident from the data, such carriers have unwittingly traveled far and wide, and formed clusters of infections in the new areas. Modeling such cases is difficult, but it may be reasonable to assume that *İ ∼ I*^*ζ*^ with *ζ <* 1, rather than *İ ∼ βI* (see next paragraph). Considering strong similarities between the rumor spreading and epidemics [5], the aforementioned long-distance travels and power-law regimes may also play a major role in rumor spreading. Note that the social media and internet provide means for fast transmission of rumor.

The aforementioned power-law growth of epidemic appears to have similarities with *turbulent diffusion* or *Taylor diffusion*, which is faster than molecular diffusion [18–22]. In turbulent diffusion, the separation between two particles, *D*(*t*), increases as *t*^3*/*2^, and 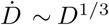 *∼ D*^1*/*3.^ The relative velocity between the particles, 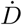, increases with time because larger eddies have larger speeds. This feature has a qualitative resemblance with aforementioned long-distance travels by asymptotic carriers.

There is possibly another connection of COVID-19 epidemic with turbulence and critical phenomena. In early stages, the epidemic spreads via contacts between infected and susceptible individuals. However, once the epidemic has spread widely, then indirect transmissions— contacts with infected surfaces, public transport, air— begin to play an important role in the epidemic growth. Such transmissions are referred to as *community spread or transmission*. This is analogous to interactions among clusters of molecules in phase transition, and those among large fluid vortices in turbulence. Such interactions are responsible for the dynamic scaling in phase transition, and for the aforementioned turbulence diffusion [23, 24, 18–22]. Super spreading of COVID-19 and the powerlaw regime of *I*(*t*) may be connected to the above phenomena. Note however that community spread could also contribute in the exponential growth phase; the two exponential regimes in Fig. 1 may be due to these reasons. These issues need further exploration.

The *I*(*t*) plots of Fig. 1 exhibit different values for the growth rate *β* and the power law exponent *n*. These constants depend on various factors, such as immunity levels of the population, climate conditions, extent of lockdowns and social distancing, etc. South Korea and China exhibit power law growths of *t*^3^ and *t*^2^ respectively, from which we may infer that a stricter lockdown may result in a relatively lower power-law exponent. The second exponential regime for USA, France, Germany, and Spain, as well as stretched power law regimes for Italy may be due to delay in the lockdowns. For India, for around a week, the exponent appears to be unity, but it may be too early to deduce anything from it. We also believe that a careful study of the *I*(*t*) curves may help in forecasting when the epidemic curve will flatten.

Now we summarize our findings. The COVID-19 real-time data of infected individuals, *I*(*t*), contains useful information that may help forecast the development of the epidemic. We conjecture that the power law growth of *I*(*t*) may be due to the epidemic transmission by asymptomatic carriers traveling long distances, and/or due to community spread.

## Data Availability

We used the data given publicly on WorldOMeter, URL https://www.worldometers.info/ coronavirus/.

## Acknowledgements

The authors thank Shashwat Bhattacharya help in early works. We also thank Shayak Bhattacharya, Prateek Sharma, and Anurag Gupta for useful discussions. Ali Asad is supported by Indo-French (CEFIPRA) project 6104-1, and Soumyadeep Chatterjee is supported by INSPIRE fellowship (IF180094) of Department of Science & Technology, India.

